# Intention of health care workers to accept COVID-19 vaccination and related factors: a systematic review and meta-analysis

**DOI:** 10.1101/2020.12.08.20246041

**Authors:** Petros Galanis, Irene Vraka, Despoina Fragkou, Angeliki Bilali, Daphne Kaitelidou

**Author notes:** Corresponding author: Petros Galanis, Assistant Professor, Faculty of Nursing, Center for Health Services Management and Evaluation, National and Kapodistrian University of Athens, 123 Papadiamantopoulou street, zip code: GR-11527, Athens, Greece.

## Abstract

Considering medical and economic burden of the coronavirus disease 2019 (COVID-19), a high COVID-19 vaccination coverage among health care workers (HCWs) is an urgent need. The aim of this systematic review and meta-analysis was to estimate the intention of HCWs to accept COVID-19 vaccination and to find out related factors. We searched PubMed, Medline, Scopus, Web of Science, ProQuest, CINAHL and medRxiv until July 14, 2021. The heterogeneity between results was very high and thus we applied a random effect model to estimate pooled effects. We performed subgroup and meta-regression analysis to identify possible resources of heterogeneity. Twenty four studies, including 39,617 HCWs met the inclusion criteria. The overall proportion of HCWs that intend to accept COVID-19 vaccination was 63.5% (95% confidence interval: 56.5-70.2%) with a wide range among studies from 27.7% to 90.1%. The following factors were associated with increased HCWs’ willingness to get vaccinated against COVID-19: male gender, older age, white HCWs, physician profession, higher education level, comorbidity among HCWs, seasonal influenza vaccination, stronger vaccine confidence, positive attitude towards a COVID-19 vaccine, fear about COVID-19, individual perceived risk about COVID-19, and contact with suspected or confirmed COVID-19 patients. The reluctance of HCWs to vaccinate against COVID-19 could diminish the trust of individuals and trigger a ripple effect in the general public. Since vaccination is a complex behavior, understanding the way that HCWs take the decision to accept or not COVID-19 vaccination will give us the opportunity to develop the appropriate interventions to increase COVID-19 vaccination uptake.

**Key Messages:**

- The overall proportion of health care workers that intent to accept COVID-19 vaccination was moderate.
- Several factors affect health care workers’ willingness to get vaccinated against COVID-19.
- COVID-19 vaccine hesitancy among health care workers should be eliminated to inspire the general public towards a positive attitude regarding a novel COVID-19 vaccine.

## 1. Introduction

Coronavirus disease 2019 (COVID-19) pandemic causes a substantial number of deaths and has a tremendous impact on the world economy [1, 2]. Globally, as of 15 July 2021, there have been more than 187 million cases of COVID-19 and more than 4 million deaths [3].

Seasonal influenza vaccination among health care workers (HCWs) is an effective infection control measure in health care settings [4, 5]. Also, the importance of HCWs vaccination against H1N1 during the 2009/2010 influenza pandemic has already been reported [6, 7]. Seasonal influenza immunization is a priority in countries with a high proportion of elderly [8–10]. During the COVID-19 pandemic, the World Health Organization (WHO) and the Centers for Disease Control and Prevention (CDC) have identified HCWs as a population with significantly elevated risk of being infected from the severe acute respiratory syndrome coronavirus 2 (SARS-CoV-2). Thus, there is a recommendation for the rapid and prioritized vaccination of HCWs against COVID-19 to protect them and the public health [11–13].

HCWs’ vaccination against infectious diseases is of utmost importance to prevent the spread of viruses, especially in health care facilities with patients. A great number of studies have already addressed the factors that influence vaccines’ acceptance by HCWs [14–19]. Several factors are identified in systematic reviews and meta-analyses such as desire for self-protection, desire to prevent illness in family or friends, perceived severity and risk of the disease, perceived safety and effectiveness of vaccination, more favorable attitudes toward vaccination, etc.

COVID-19 vaccination acceptance among HCWs is essential to decrease the spread of the SARS-CoV-2 and to protect public health. Moreover, HCWs could inform and educate people about COVID-19 vaccines building confidence in vaccines and promoting acceptance. To date, no systematic review and meta-analysis has investigated the willingness of HCWs to accept COVID-19 vaccination. Thus, we performed a systematic review and meta-analysis to estimate the intention of HCWs to accept COVID-19 vaccination and to find out related factors.

## 2. Materials and methods

### 2.1. Data sources and strategy

We followed the Preferred Reporting Items for Systematic Reviews and Meta-Analysis (PRISMA) guidelines for this systematic review and meta-analysis [20]. We searched PubMed, Medline, Scopus, Web of Science, ProQuest, CINAHL and pre-print services (medRxiv) for articles published from January 1, 2020 to July 14, 2021. Through the databases, in the query box we used the following strategy in all fields: (((“health care worker•” OR “healthcare worker•” OR “healthcare personnel” OR “health care personnel” OR “health personnel” OR “health care professional•” OR “healthcare professional•” OR HCWS OR HCW OR HCPS OR HCP OR staff OR “nursing staff” OR employee• OR professional• OR personnel OR worker• OR doctor• OR physician• OR clinician• OR nurs• OR midwives OR midwife• OR paramedic• OR hospital• OR practitioner•) AND (vaccin•)) AND (intent• OR willing• OR hesitancy)) AND (COVID-19 OR COVID19 OR COVID OR SARS-CoV• OR “Severe Acute Respiratory Syndrome Coronavirus•” OR coronavirus•). Also, we examined reference lists of all relevant articles that we found during the search process. Finally, we removed duplicates.

### 2.2. Selection and eligibility criteria

Study selection was performed by two independent reviewers, while a third, senior reviewer resolved the discrepancies. Firstly, we screened title, then abstract of the records and finally the full-text. We applied the following inclusion criteria: studies examining HCWs’ intention to accept COVID-19 vaccination and related factors; studies that were written in English; studies included all types of HCWs working in clinical settings. On the other hand, we excluded qualitative studies, reviews, case reports, protocols, editorials, and letters to the Editor. Also, we excluded studies including students of health sciences, retired HCWs, patients, and general population.

### 2.3. Data extraction and quality assessment

We extracted the following data from each study: authors, location, sample size, age, gender, study design, sampling method, assessment of intention to accept COVID-19 vaccination, response rate, data collection time, type of publication (journal or pre-print service), number of HCWs with intention to accept COVID-19 vaccination, type of occupation (physicians, nurses, assistant nurses, paramedical staff, etc), factors related with intention to accept COVID-19 vaccination, and the level of analysis (univariate or multivariable). Assessment of intention to accept COVID-19 vaccination was referred to vaccine acceptance (e.g., binary yes/no answer, five or eleven point Likert-type scale). Perceived risk of COVID-19 is a combination of subjective perception of disease severity and susceptibility [21]. Fear of COVID-19 among HCWs mainly includes fear of getting sick with the disease and fear of infecting patients, family members, and friends [22]. Attitudes toward COVID-19 vaccination are defined as expressions of hesitancy or support measuring usually in Likert scales [23].

Two independent reviewers used the Joanna Briggs Institute critical appraisal tools to assess quality of studies (poor, moderate or good quality). An 8-point scale is used for cross-sectional studies with a score of ≤3 indicates poor quality, a score of 4-6 points indicates moderate quality, and a score of 7-8 points indicates good quality [24]. The Joanna Briggs Institute critical appraisal tool for cross-sectional studies includes eight different assessment domains e.g., inclusion criteria for the sample, detailed description of the settings, exposure and outcome measurement, identification of confounding factors and strategies to eliminate them, and statistical analysis.

### 2.4. Statistical Analysis

For each study we divided the number of HCWs with intention to accept COVID-19 vaccination with the sample size to calculate the proportion of HCWs with intention to accept vaccination and the relative 95% confidence interval (CI). Then, we transformed the proportions with the Freeman-Tukey Double Arcsine method before pooling [25]. Studies that used Likert scales to assess the intention to accept COVID-19 vaccination considered the answers “agree”/“strongly agree” as a positive answer. We used the I^2^ and Hedges Q statistics to assess between-studies heterogeneity. I^2^ values higher than 75% indicate high heterogeneity and a p-value<0.1 for the Hedges Q statistic indicates statistically significant heterogeneity [26]. The heterogeneity between results was very high and thus we applied a random effect model to estimate pooled effects [26]. We considered sample size, age, gender, response rate, data collection time, publication type (journal or pre-print service), type of occupation, studies quality, and the continent that studies were conducted as pre-specified sources of heterogeneity. Due to the limited variability of data in some variables, we decided to perform subgroup analysis for publication type, studies quality, and the continent that studies were conducted and meta-regression analysis for sample size, gender distribution, and data collection time as the independent variables. We conducted a leave-one-out sensitivity analysis to determine the influence of each study on the overall effect. This type of analysis performs sequent meta-analyses by leaving out exactly one study at each meta-analysis. In that case, we can investigate the way that each study affects the overall effect size estimate identifying influential studies. The Egger’s test was used to assess the publication bias with a P-value<0.05 indicating publication bias [27]. We did not perform meta-analysis for the factors related with intention of HCWs to accept COVID-19 vaccination since the data were highly heterogeneous and limited. We used OpenMeta[Analyst] for the meta-analysis [28].

## 3. Results

### 3.1. Identification and selection of studies

Flowchart of the literature search according to PRISMA guidelines is presented in Figure 1. Initially, we identified 3022 potential records through electronic databases and 730 duplicates were removed. After the screening of the titles and abstracts, we removed 2114 records and we added one more record found by the reference lists scanning. We included 24 studies in this systematic review and meta-analysis that met our inclusion criteria.

**Figure 1.**
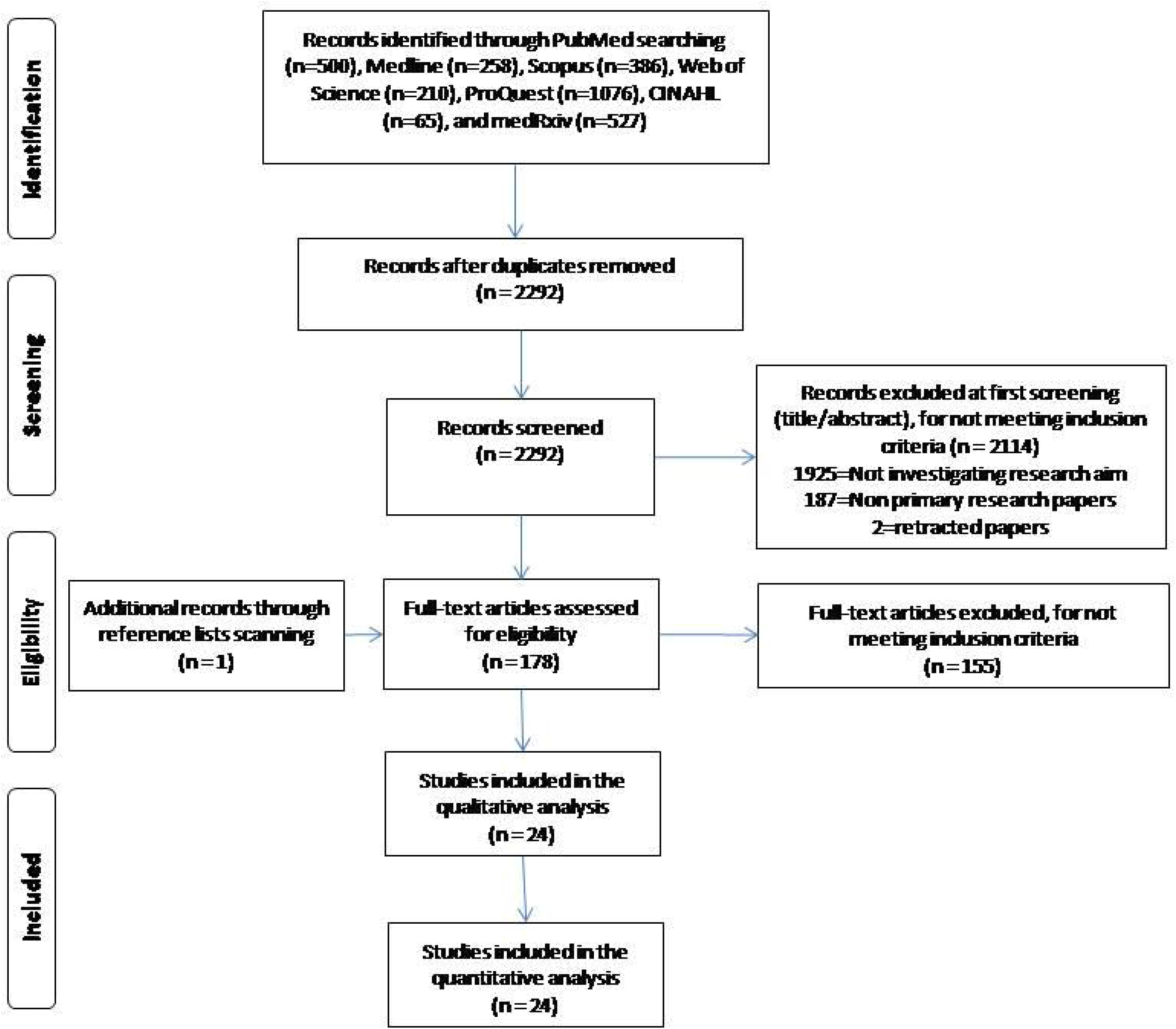
Flowchart of the literature search according to the Preferred Reporting Items for Systematic Reviews and Meta-Analysis.

### 3.2. Characteristics of the studies

Main characteristics of the 24 studies included in this review are presented in Table 1. A total of 39,617 HCWs were included in this systematic review with a minimum of 208 HCWs [29] and a maximum of 12,034 HCWs [30] among studies. Ten studies were conducted in Asia (China, Turkey, Kingdom of Saudi Arabia, Vietnam, Kuwait and Hong Kong) [31–40], six studies in North America (USA and Canada) [30, 41–45], four studies in Europe (France, Germany and Greece) [46–49], three studies in Africa (Democratic Republic of the Congo, Eastern Cape and Zambia) [29, 50, 51], and one study was multicenter (France, Belgium and Canada) [52]. Females were more in 19 studies [30–37, 39–47, 51, 52], while males were more in four studies [29, 38, 49, 50]. All studies were cross-sectional, while 23 studies used convenience sampling method and one used snowball sampling method [44]. Nineteen studies were published in journals [30, 31, 34, 36–40, 42–52] and fine studies in pre-print services [29, 32, 33, 35, 41]. One study did not report data regarding age [48], one regarding gender distribution [48], and 12 regarding response rate [29, 34, 37, 39, 40, 44–46, 48–51]. Ten studies used a yes/no answer to assess intention of HCWs to accept COVID-19 vaccination [30, 35, 36, 39, 41, 43, 45, 46, 50, 51], nine studies used a yes/no/uncertain answer [29, 31, 32, 34, 37, 38, 40, 44, 52], and five studies used Likert-type scales [33, 42, 47–49].

**Table 1.** Main characteristics of the studies included in this systematic review.

| Reference | Location | Sample size (n) | Age, mean (SD) | Females (%) | Study design | Sampling method | Assessment of intention to accept COVID-19 vaccination | Response rate (%) | Data collection time | Publication in |
| --- | --- | --- | --- | --- | --- | --- | --- | --- | --- | --- |
| Gagneux-Brunon et al. (2020)[46] | France | 2047 | <30 years: 22.7%; 30-49: 47.3%; 50-64: 26.8%; >64: 3.1% | 74 | Cross-sectional | Convenience sampling | Yes/no answer | NR | March 26 to July 2, 2020 | Journal |
| Nzaji et al. (2020)[50] | Democratic Republic of the Congo | 613 | 40.3 (11.7) | 49.1 | Cross-sectional | Convenience sampling | Yes/no answer | NR | March 01 to April 30, 2020 | Journal |
| Papagiannis et al. (2020)[47] | Greece | 461 | 44.2 (10.8) | 74 | Cross-sectional | Convenience sampling | Five point Likert-type scale (fully disagree to fully agree) | 92.2 | February 10-25, 2020 | Journal |
| Wang et al. (2020)[31] | Hong Kong | 806 | <30 years: 21.6%;<br>30-39: 31.1%; 40-49: 27.1%; >49: 20.2% | 87.5 | Cross-sectional | Convenience sampling | Yes/no/uncertain answer | 5.2 | February 26 to March 21, 2020 | Journal |
| Detoc et al. (2020)[48] | France | 1421 | NR | NR | Cross-sectional | Convenience sampling | Five point Likert-type scale (definitely no to certainly yes) | NR | March 26 to April 20, 2020 | Journal |
| Fu et al. (2020)[32] | China | 352 | <30 years: 36.9%;<br>30-39: 31.8%; 40-49: 22.2%; >49: 9.1% | 58.8 | Cross-sectional | Convenience sampling | Yes/no/uncertain answer | 96.2 | March 17-18, 2020 | Pre-print service |
| Kwok et al. (2020)[33] | Hong Kong | 1205 | 40.8 (10.5) | 89.7 | Cross-sectional | Convenience sampling | Eleven-point Likert-type scale (0=definitely no, 10=definitely yes) | 78.9 | March 16 to April 29, 2020 | Pre-print service |
| Chawe et al. (2020)[29] | Zambia | 208 | <30 years: 50%; 30-39: 38%; >39: 12% | 41.8 | Cross-sectional | Convenience sampling | Yes/no/uncertain answer | NR | June 10-29, 2020 | Pre-print service |
| Gadoth et al. (2020)[41] | USA | 609 | <30 years: 14.6%; 30-39: 37.8%; 40-49: 22.3%; >49: 21.4% | 68.8 | Cross-sectional | Convenience sampling | Yes/no answer | 55.7 | September 24 to October 16, 2020 | Pre-print service |
| Verger et al. (2021)[52] | France, Belgium, Canada | 2678 | 18-39 years: 34.4%; 40-59 years: 46.7%; >59 | 69.3 | Cross-sectional | Convenience sampling | Yes/no/uncertain answer | 17.6 | October to November, 2020 | Journal |
|  |  |  | years: 18.9% |  |  |  |  |  |  |  |
| Shaw et al. (2021)[42] | USA | 5287 | 42.5 (13.6) | 72.7 | Cross-sectional | Convenience sampling | Five point Likert-type scale (fully disagree to fully agree) | 55.0 | November 23 to December 5, 2020 | Journal |
| Unroe et al. (2021)[43] | USA | 8243 | 18-24 years: 12.7%; 25-40 years: 37%; 41-60 years: 38.9%; >60 years: 11.4% | 86.8 | Cross-sectional | Convenience sampling | Yes/no answer | 33 | November 14-17, 2020 | Journal |
| Shekhar et al. (2021)[44] | USA | 3479 | <40 years: 54%; ≥40 years: 46% | 75 | Cross-sectional | Snowball sampling | Yes/no/uncertain answer | NR | October 7 to November 9, 2020 | Journal |
| Kose et al. (2021)[34] | Turkey | 1138 | <25 years: 89.6%; | 72.5 | Cross- | Convenience | Yes/no/uncertain | NR | September 17-20, | Journal |
|  |  |  | ≥25 years: 10.4% |  | sectional | sampling | answer |  | 2020 |  |
| Kuter et al. (2021)[30] | USA | 12,034 | <40 years: 51%;<br>40-64 years:<br>39.2%; >64 years:<br>3.6% | 71.7 | Cross-sectional | Convenience<br>sampling | Yes/no answer | 34.5 | November 13 to<br>December 6,<br>2020 | Journal |
| Barry et al. (2020)[35] | Kingdom of<br>Saudi Arabia | 1512 | 21-30 years:<br>25.5%; 31-40<br>years: 44.8%; 41-50 years: 19.7%;<br>>50 years: 10.1% | 62.4 | Cross-sectional | Convenience<br>sampling | Yes/no answer | 75.3 | November 4-14,<br>2020 | Pre-print<br>service |
| Huynh et al. (2021)[36] | Vietnam | 410 | 39.3 (9.3) | 68.8 | Cross-sectional | Convenience<br>sampling | Yes/no answer | 48.0 | January to<br>February, 2021 | Journal |
| Kaplan et al. (2021)[37] | Turkey | 1574 | 39.4 (10.8) | 58.8 | Cross-sectional | Convenience<br>sampling | Yes/no/uncertain<br>answer | NR | December 25-31,<br>2020 | Journal |
| Qattan et al. (2021)[38] | Kingdom of Saudi Arabia | 673 | <39 years: 67.1%;<br>≥39 years: 32.9% | 39.8 | Cross-sectional | Snowball sampling | Yes/no/uncertain answer | 91.4 | December 8-14, 2020 | Journal |
| Nohl et al. (2021)[49] | Germany | 1296 | <40 years: 63.8%;<br>≥40 years: 36.2% | 21.8 | Cross-sectional | Convenience sampling | Five point Likert-type scale (fully disagree to fully agree) | NR | December 4, 2020 to January 15, 2021 | Journal |
| Dzieciolowska et al. (2021)[45] | Canada | 2761 | 44.0 (6.5) | 72.2 | Cross-sectional | Convenience sampling | Yes/no answer | NR | December 15-28, 2020 | Journal |
| Sun et al. (2021)[39] | China | 505 | 32.4 (8.9) | 77.4 | Cross-sectional | Convenience sampling | Yes/no answer | NR | January 4-6, 2021 | Journal |
| Al-Sanafi & Sallam (2021)[40] | Kuwait | 1019 | 34 (9.7) | 61.4 | Cross-sectional | Convenience sampling | Yes/no/uncertain answer | NR | March 18-29, 2021 | Journal |
| Adeniyi et al. (2021)[51] | Eastern | 1308 | 26-55 years: 79.1%; >55 years: | 81.5 | Cross- | Convenience | Yes/no answer | NR | November to | Journal |
|  | Cape |  | 20.9% |  | sectional | sampling |  |  | December, 2020 |  |

NR: not reported

Intention of HCWs to accept vaccination and study population in the studies included in this systematic review are presented in Table 2. Intention ranged from 27.7% [50] to 90.1% [51]. Percentage of physicians that participated in studies ranged from 12.1% [34] to 60.6% [52], while percentage of nurses ranged from 12.5% [40] to 100% [31, 33]. Four studies did not report detailed data regarding study population [32, 38, 48, 49].

**Table 2.**
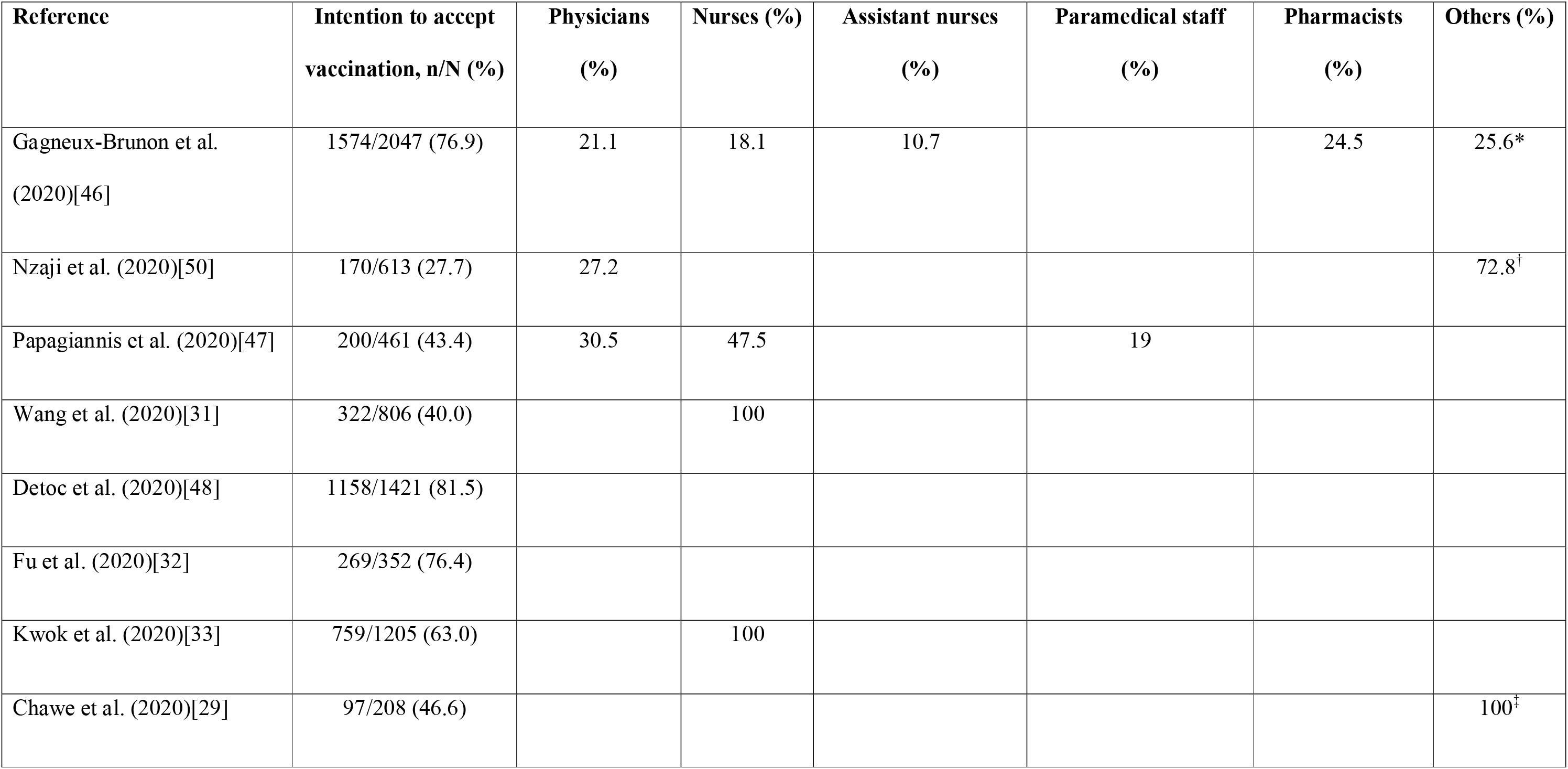
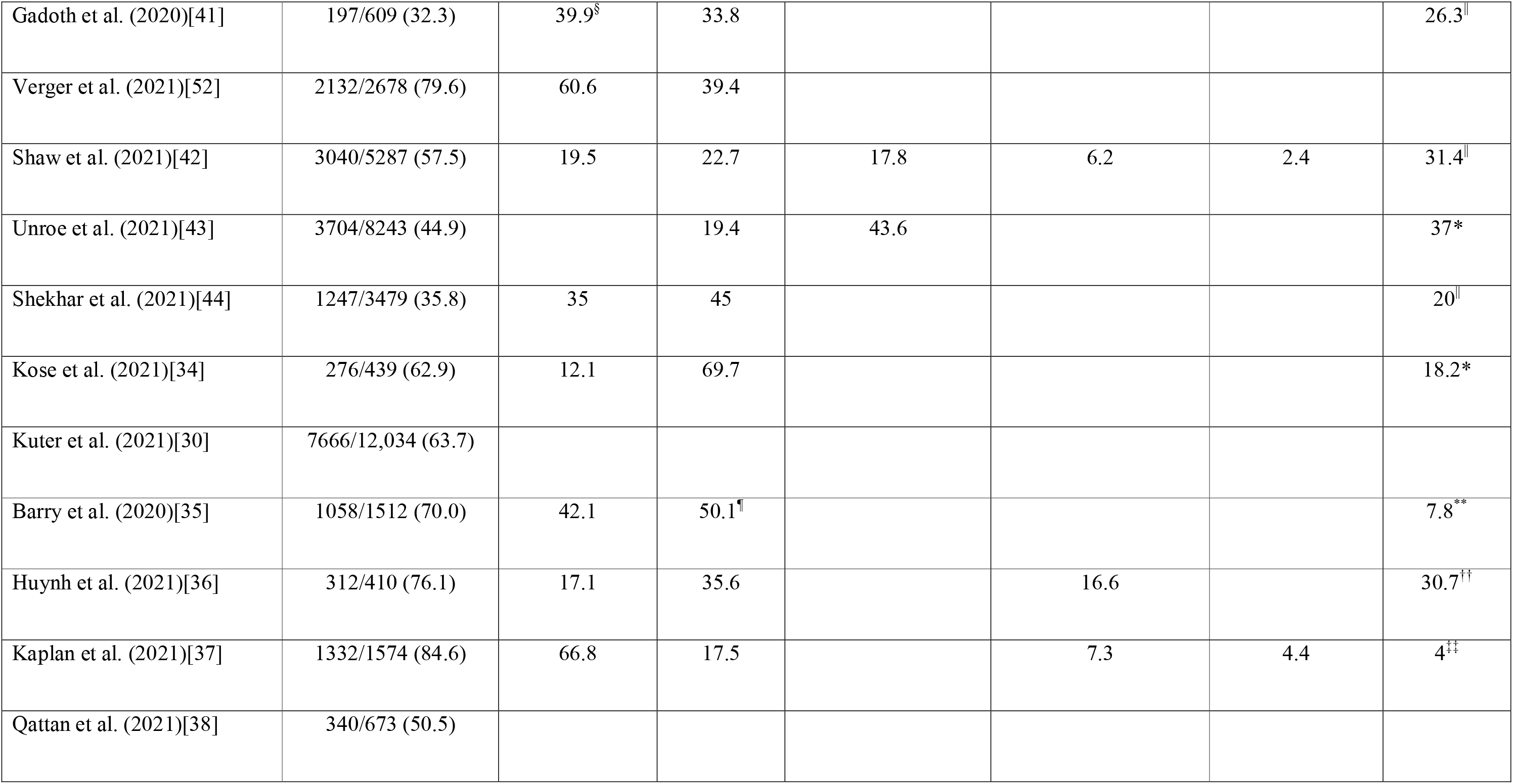
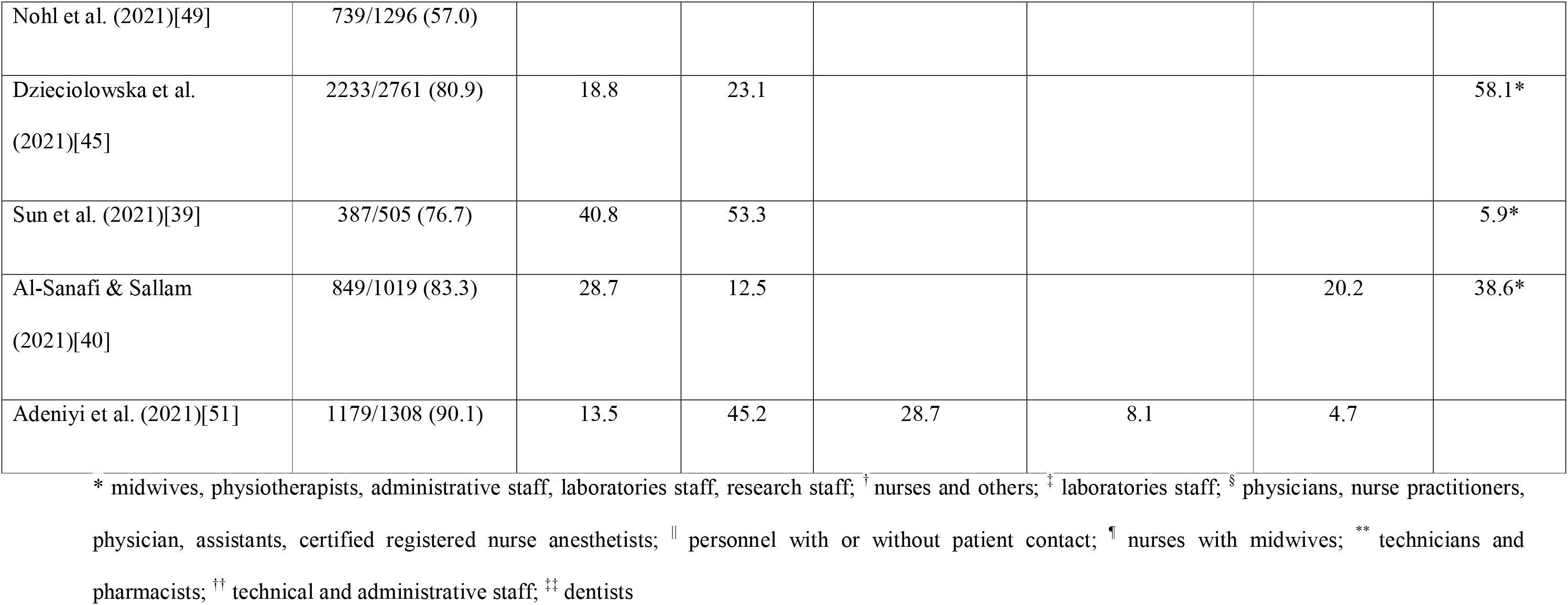
Intention of health care workers to accept vaccination and study population in the studies included in this systematic review.

### 3.3. Quality assessment

Quality assessment of cross-sectional studies included in this review is shown in Table 3. Quality was moderate in six studies [29, 32, 34, 42, 47, 48] and good in 18 studies [30, 31, 33, 35, 37–41, 43–46, 49–52].

**Table 3.** Quality of cross-sectional studies included in this systematic review.

|  | Gagneux-Brunon et<br>al.<br>(2020)[46] | Nzaji et<br>al.<br>(2020)[50] | Papagiannis<br>et al.<br>(2020)[47] | Wang et<br>al.<br>(2020)[31] | Detoc et<br>al.<br>(2020)[48] | Fu et al.<br>(2020)[32] | Kwok et<br>al.<br>(2020)[33] | Chawe et<br>al.<br>(2020)[29] | Gadoth et<br>al.<br>(2020)[41] | Verger et<br>al.<br>(2021)[52] |
| --- | --- | --- | --- | --- | --- | --- | --- | --- | --- | --- |
| 1. Were the criteria for inclusion in the sample clearly defined? | √ | √ | √ | √ |  | √ | √ |  | √ | √ |
| 2. Were the study subjects and the setting described in detail? | √ | √ | √ | √ |  |  | √ | √ | √ | √ |
| 3. Was the exposure measured in a valid and reliable way? | √ | √ | √ | √ | √ | √ | √ | √ | √ | √ |
| 4. Were objective, standard criteria used for measurement of the condition? | √ | √ | √ | √ | √ | √ | √ | √ | √ | √ |
| 5. Were confounding factors identified? | √ | √ |  | √ |  |  | √ |  | √ | √ |
| 6. Were strategies to deal with confounding factors stated? | √ | √ |  | √ |  |  | √ |  | √ | √ |
| 7. Were the outcomes measured in a valid and reliable way? | √ | √ | √ | √ | √ | √ | √ | √ | √ | √ |
| 8. Was appropriate statistical analysis used? | √ | √ | √ | √ | √ | √ | √ | √ | √ | √ |
| **Total quality** | Good | Good | Moderate | Good | Moderate | Moderate | Good | Moderate | Good | Good |

**Table 3 (continued).** Quality of cross-sectional studies included in this systematic review.
|  | Shaw et al.<br>(2021)[42] | Unroe et al.<br>(2021)[43] | Shekhar et al.<br>(2021)[44] | Kose et al.<br>(2021)[34] | Kuter et al.<br>(2021)[30] | Barry et al.<br>(2020)[35] | Huynh et al.<br>(2021)[36] | Kaplan et al.<br>(2021)[37] | Qattan et al.<br>(2021)[38] | Nohl et al.<br>(2021)[49] |
| --- | --- | --- | --- | --- | --- | --- | --- | --- | --- | --- |
| 1. Were the criteria for inclusion in the sample clearly defined? | √ | √ | √ |  | √ | √ |  |  | √ |  |
| 2. Were the study subjects and the setting described in detail? | √ | √ | √ | √ | √ | √ | √ | √ | √ | √ |
| 3. Was the exposure measured in a valid and reliable way? | √ | √ | √ |  | √ | √ | √ | √ | √ | √ |
| 4. Were objective, standard criteria used for measurement of the condition? | √ | √ | √ | √ | √ | √ | √ | √ | √ | √ |
| 5. Were confounding factors identified? |  | √ | √ |  | √ | √ | √ | √ | √ | √ |
| 6. Were strategies to deal with confounding factors stated? |  | √ | √ |  | √ | √ | √ | √ | √ | √ |
| 7. Were the outcomes measured in a valid and reliable way? | √ | √ | √ | √ | √ | √ | √ | √ | √ | √ |
| 8. Was appropriate statistical analysis used? | √ | √ | √ | √ | √ | √ | √ | √ | √ | √ |
| **Total quality** | Moderate | Good | Good | Moderate | Good | Good | Good | Good | Good | Good |

**Table 3 (continued).** Quality of cross-sectional studies included in this systematic review.
|  | Dzieciolowska<br>et al.<br>(2021)[45] | Sun et al.<br>(2021)[39] | Al-Sanafi & Sallam<br>(2021)[40] | Adeniyi et al.<br>(2021)[51] |
| --- | --- | --- | --- | --- |
| 1. Were the criteria for inclusion in the sample clearly defined? | √ |  | √ | √ |
| 2. Were the study subjects and the setting described in detail? | √ | √ | √ | √ |
| 3. Was the exposure measured in a valid and reliable way? | √ | √ | √ | √ |
| 4. Were objective, standard criteria used for measurement of the condition? | √ | √ | √ | √ |
| 5. Were confounding factors identified? | √ | √ | √ | √ |
| 6. Were strategies to deal with confounding factors stated? | √ | √ | √ | √ |
| 7. Were the outcomes measured in a valid and reliable way? | √ | √ | √ | √ |
| 8. Was appropriate statistical analysis used? | √ | √ | √ | √ |
| **Total quality** | Good | Good | Good | Good |

### 3.4. Meta-analysis

The overall proportion of HCWs that intend to accept COVID-19 vaccination was 63.5% (95% CI: 56.5-70.2%) (Figure 2). The heterogeneity between results was very high (I^2^=99.59%, p-value for the Hedges Q statistic<0.001). A leave-one-out sensitivity analysis showed that no single study had a disproportional effect on the pooled proportion, which varied between 62.1% (95% CI: 55.3-68.7%), with Adeniyi et al. [51] excluded, and 65.0% (95% CI: 58.1-71.6%), with Nzaji et al. [50] excluded (Supplementary Figure S1).

**Figure 2.**
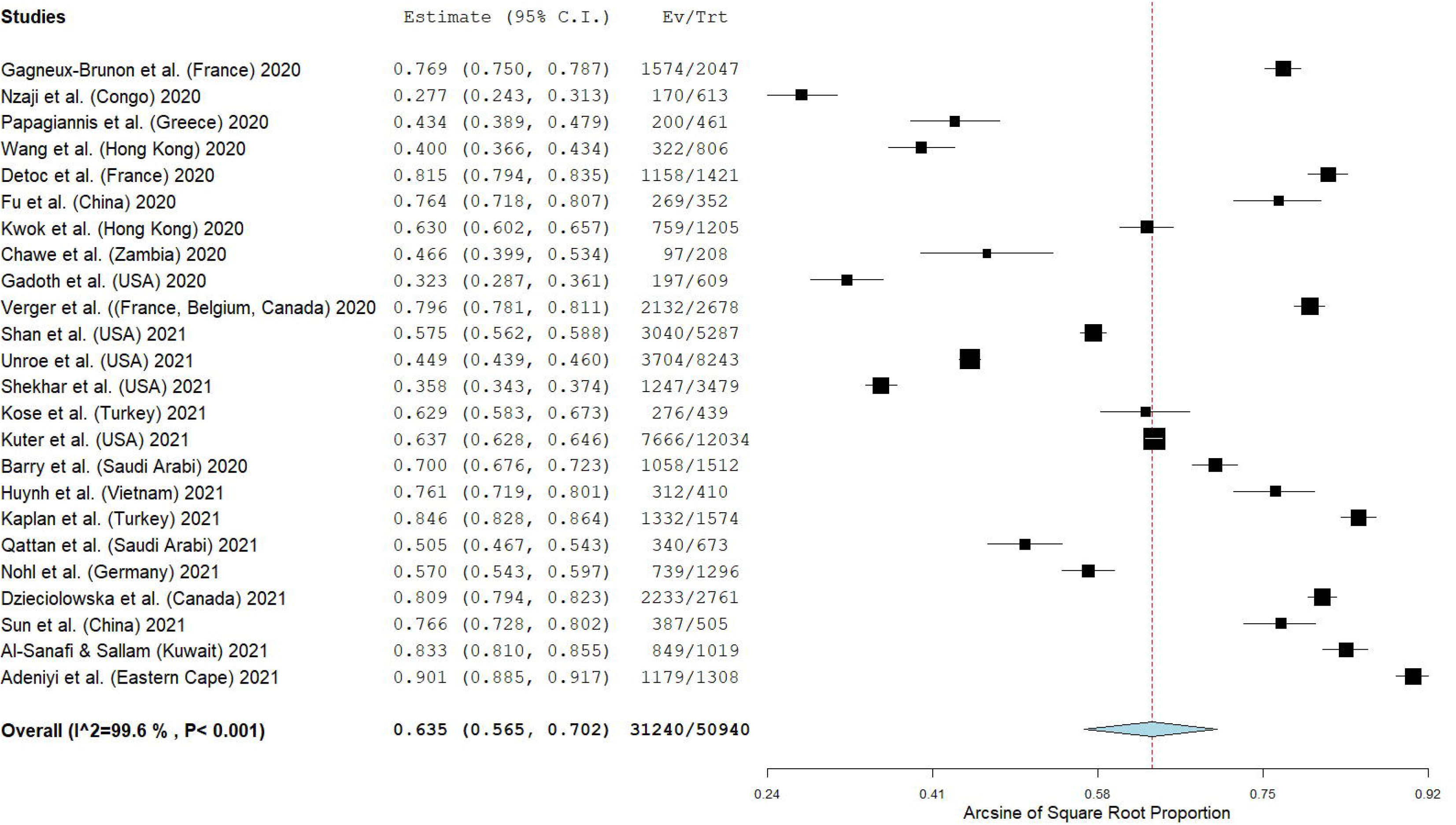
Forest plot of the proportion of HCWs that intend to accept COVID-19 vaccination.

According to subgroup analysis, the proportion of HCWs that intend to accept COVID-19 vaccination was higher for the studies that were published in journals (64.9% [95% CI: 57.0-72.4%], I^2^=99.66) than those in pre-print services (58.0% [95% CI: 43.2-72.2%], I^2^=98.75). Moreover, the proportion was almost the same for the studies with moderate quality (62.0% [95% CI: 49.5-73.8%], I^2^=98.86) and those with good quality (64.0% [95% CI: 55.4-72.1%], I^2^=99.68). The proportion of HCWs that intend to accept COVID-19 vaccination was higher in studies that were conducted in Europe (65.5% [95% CI: 50.0-79.6%], I^2^=99.22) and Asia (69.0% [95% CI: 59.4-77.9%], I^2^=98.84) compared to those in Africa (56.7% [95% CI: 12.2-95.2%], I^2^=99.77) and North America (52.9% [95% CI: 40.8-64.9%], I2=99.78). Meta-regression showed that the closer each study was performed to now, the more likely HCWs were to accept COVID-19 vaccination (coefficient=0.024, [95% CI: 0.006-0.042], p=0.008). Also, the proportion of HCWs that intend to accept COVID-19 vaccination was independent of the sample size (p=0.17), and gender distribution (p=0.15). P-value<0.05 for Egger’s test implied potential publication bias.

### 3.5. Factors related with intention of HCWs to accept COVID-19 vaccination

Twenty studies [30, 31, 33, 35–47, 49–52] investigated factors related with intention of HCWs to accept COVID-19 vaccination, while 18 studies [30, 31, 33, 35–41, 43–46, 49–52] used multivariable analysis to control confounding (Table 4). Statistically significant factors are presented in Table 4 and were discussed in the following paragraphs.

**Table 4.**
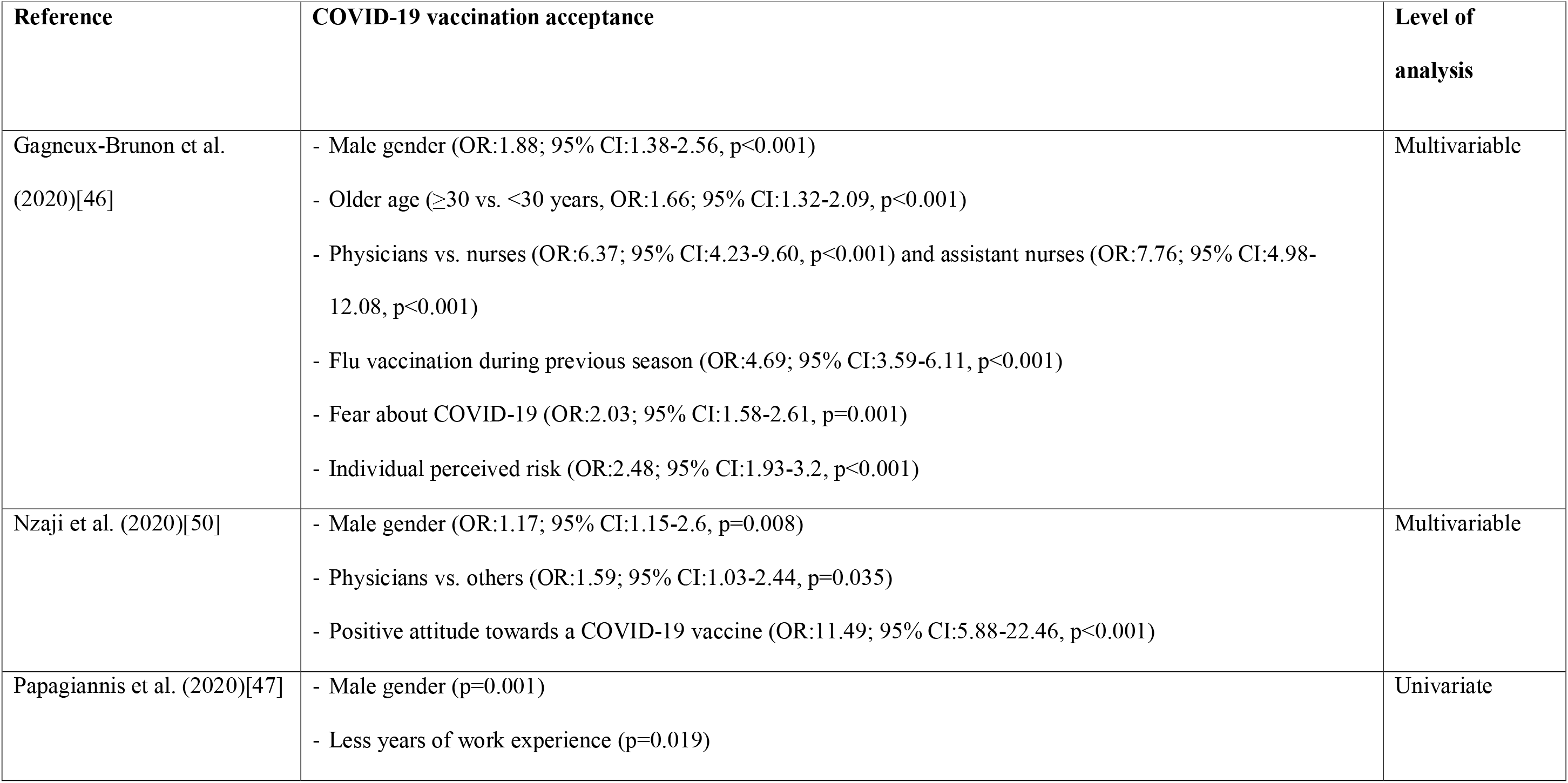
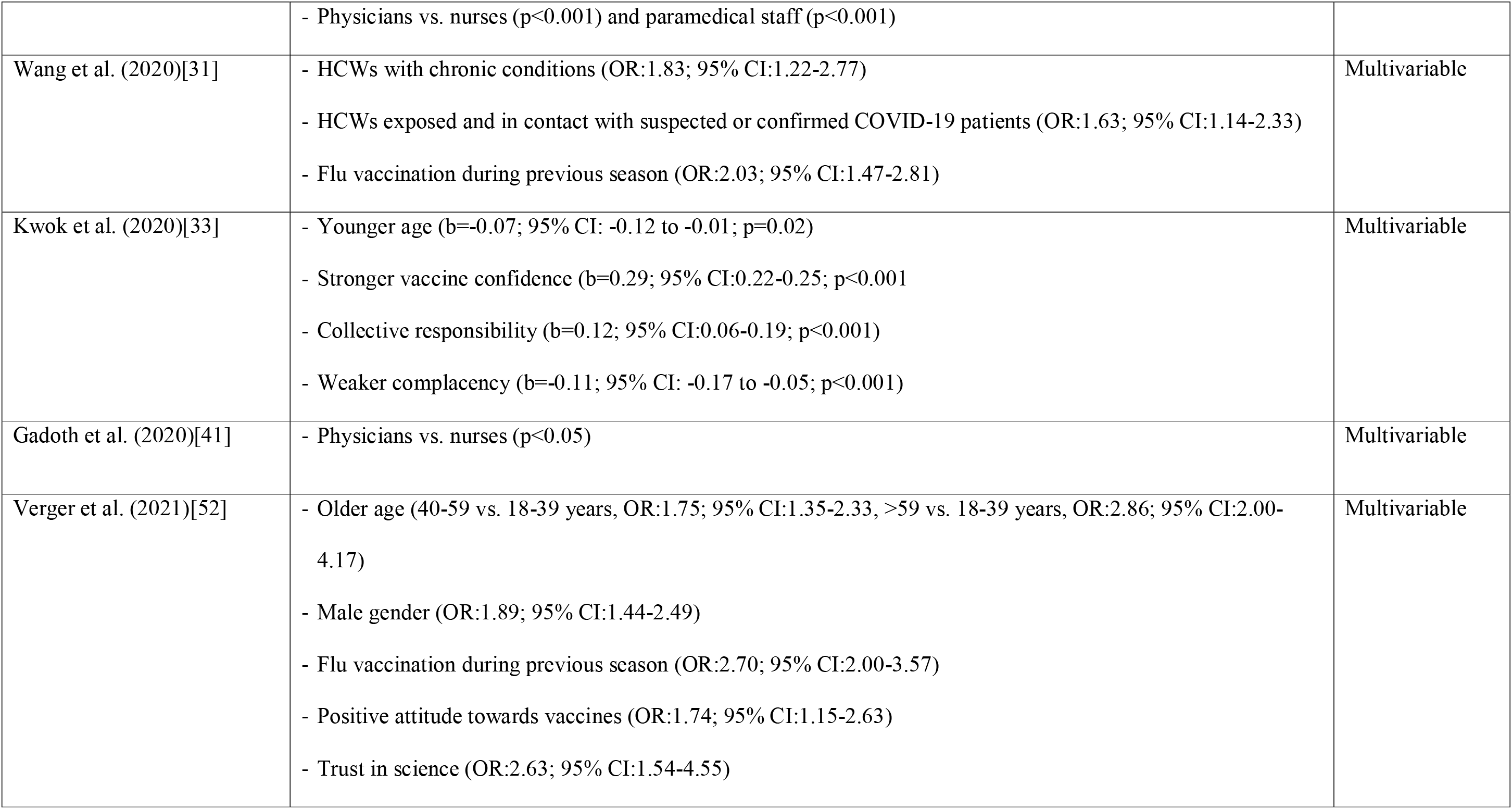
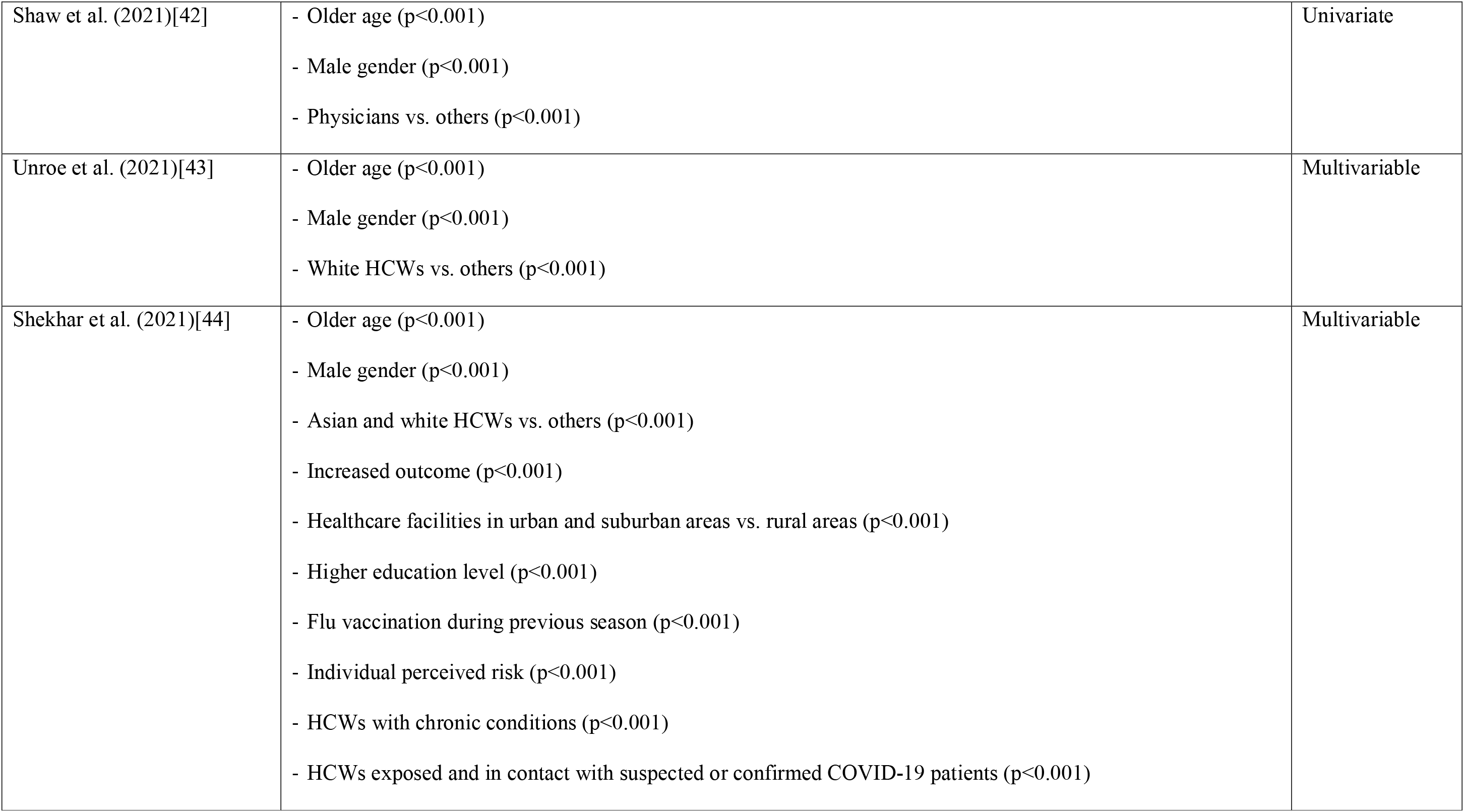
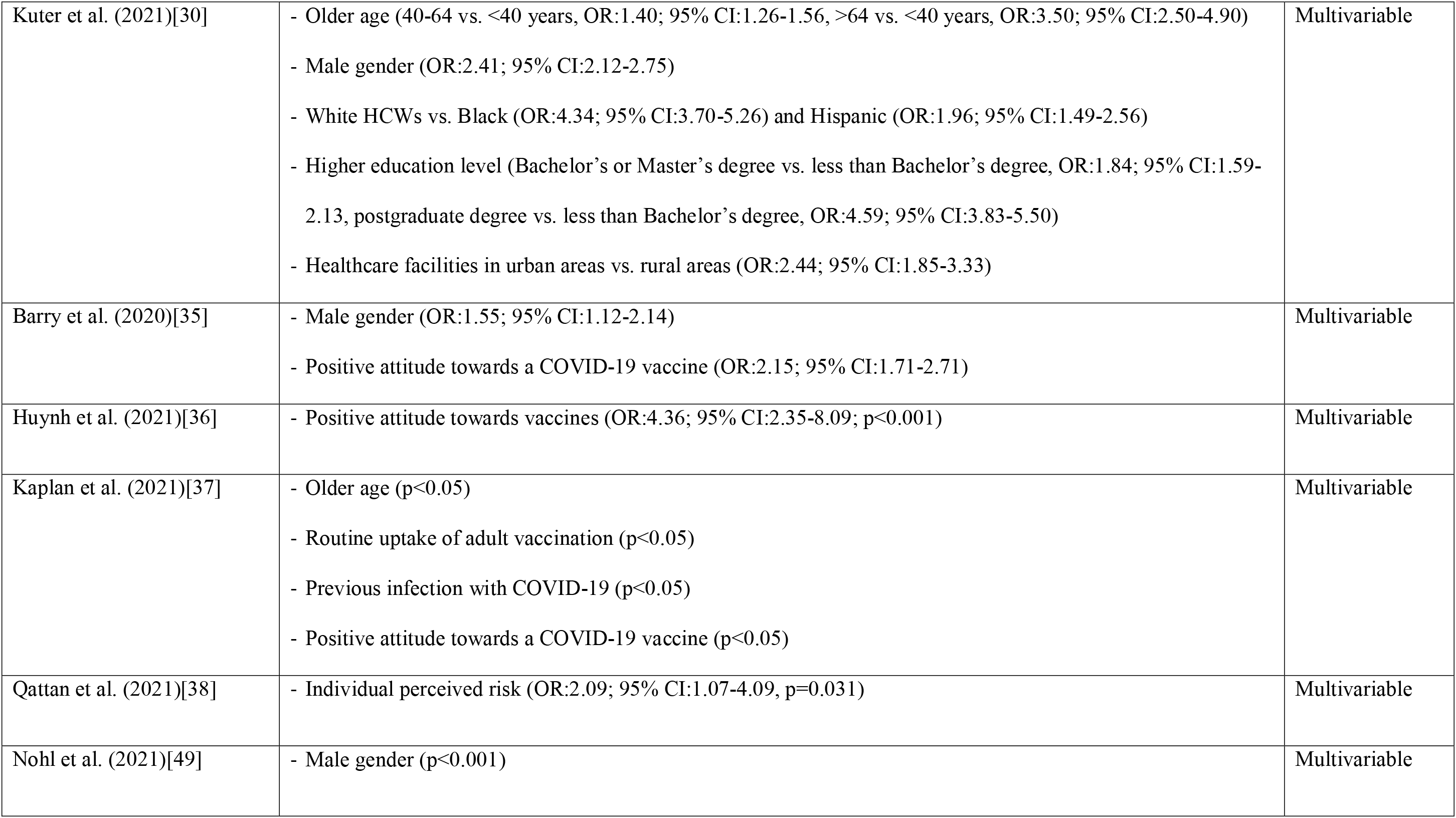
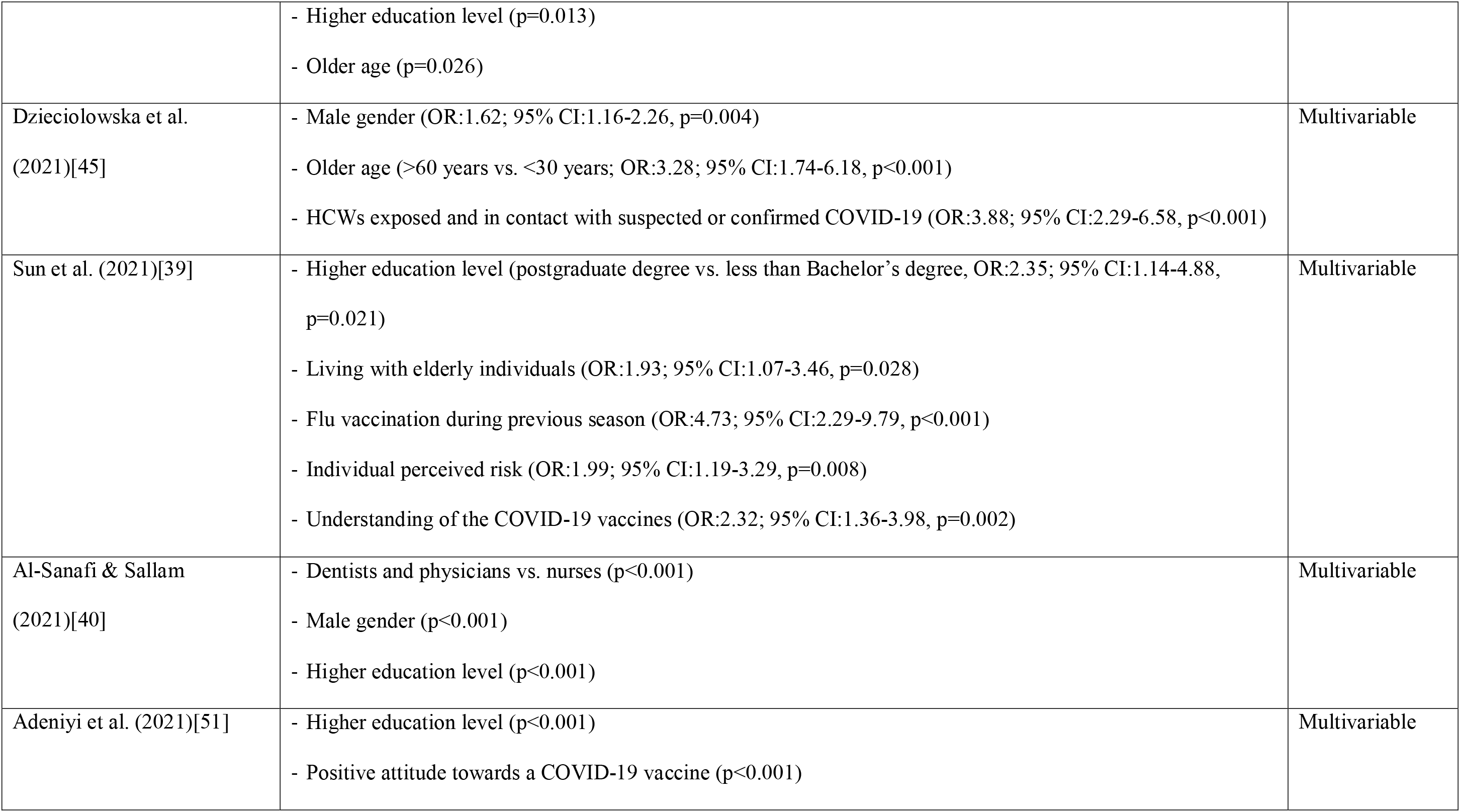
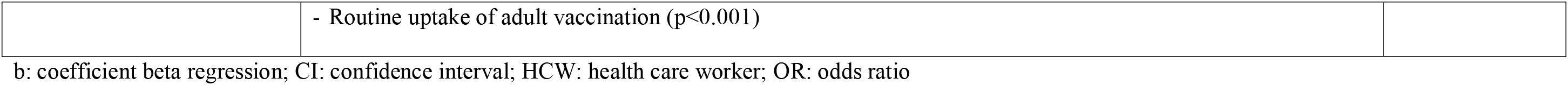
Statistically significant factors related with intention of health care workers to accept COVID-19 vaccination.

We found that several demographic characteristics were associated with COVID-19 vaccination acceptance. Profession was an important predictor since six studies [40–42, 46, 47, 50] found that physicians were more prone to get vaccinated against COVID-19 than other HCWs and especially nurses and paramedical staff. Male HCWs [30, 31, 37, 43–47, 49, 50, 52] and white HCWs [30, 43, 44] were more likely to be vaccinated. A great number of studies [30, 37, 42–46, 49, 52] found that older age was associated with an increase in COVID-19 vaccine acceptance. Higher education level [30, 39, 40, 44, 49, 51], increased outcome [44], and work in healthcare facilities in urban areas [30, 44] were related with increased COVID-19 vaccine acceptance. Also, HCWs with chronic conditions were more likely to be vaccinated against COVID-19 [31, 44].

Flu vaccination during previous season was associated with intention to accept COVID-19 vaccination [31, 37, 39, 44, 46, 51, 52]. Stronger vaccine confidence [33] and positive attitude towards a COVID-19 vaccine [35, 37, 39, 44, 50–52] increased HCWs’ willingness to get vaccinated against COVID-19. Fear about COVID-19 [46], individual perceived risk about COVID-19 [38, 39, 44, 46], and weaker complacency about the COVID-19 [33] were related with increased COVID-19 vaccination acceptance. Complacency was measured on a 7-point Likert scale (strongly disagree to strongly agree). HCWs exposed and in contact with suspected or confirmed COVID-19 patients [31, 44, 45] and those with a previous COVID-19 infection [37] were more likely to accept COVID-19 vaccine.

## 4. Discussion

To our knowledge, this is the first systematic review and meta-analysis that assesses the intention of HCWs to accept COVID-19 vaccination and related factors. Twenty-four papers met our inclusion criteria and the primary reasons that other papers were excluded from this review include irrelevant research question, study population other than HCWs, and other types of publications (e.g. qualitative studies, reviews, case reports, protocols, editorials, and letters to the Editor). We found that the proportion of HCWs that intend to accept COVID-19 vaccination was moderate (63.5%) with a wide range among studies from 27.7% to 90.1%. This moderate level of acceptance may be attributable to several reasons, e.g. inadequate knowledge among HCWs regarding COVID-19 [37, 53], negative attitude towards the disease [53, 54], and feelings of fear and anxiety during the COVID-19 pandemic [55–58]. Also, concerns raised for COVID-19 vaccination are related with inadequate knowledge about such new vaccines regarding the long term side effects, effectiveness, efficacy etc. Better knowledge of COVID-19 among HCWs affects their attitude, increases their confidence, and promotes preventive measures such as the vaccination [59–61]. According to our subgroup analysis, the proportion of HCWs that intend to accept COVID-19 vaccination was higher in studies in Europe and Asia than those in North America and Africa. This finding is in accordance with a study [62] in 10 countries in Africa, Latin America, Eastern Europe, Asia Pacific, and the Middle East where the influenza vaccination coverage rate in general population was much higher in Europe than in Asia and Africa. This difference may be attributable mainly to the fact that a national influenza vaccination policy and recommendations for seasonal influenza vaccination are standard in developed countries but this is not the case in many developing countries in Africa. Also, the availability of influenza vaccines is low in Africa [63], while the number of influenza vaccines per capita is much higher in high-income countries compared to lower and middle-income countries (median number; 139.2 vs. 6.1 per 1000 population) [64].

Τhe positive effects of the influenza vaccine in health outcomes and in financial terms are well known [10, 65–67], but the vaccination rate is low even among HCWs. A meta-analysis [19] with 45 studies in mainland China found that the influenza vaccination rate was 17.7%, 9.4%, 7.8%, and 3.5% for HCWs, general population, pregnant women, and people with chronic conditions respectively. A similar finding was found in studies in Europe (United Kingdom, Germany, France, and Spain) where HCWs received influenza vaccination more often than the general population but in low levels, ranging from 15% to 29% [68]. A meta-analysis [69] included studies in Italy found that the proportion of influenza vaccination among nurses and ancillary workers was 13.47% and 12.52% respectively. Influenza vaccination coverage is higher in the USA (80.6%) [70] and Canada (ranging from 35.5% to 51%) [71, 72], but still lower than the national Healthy People 2020 target of 90% [73].

We found a difference in intention to accept COVID-19 vaccination between the professions, with physicians most inclined to get vaccinated compared to other HCWs and especially nurses and paramedical staff. This finding is confirmed by two meta-analyses [69, 74] including studies in Italy, where the prevalence of influenza vaccination among physicians was 23.18% [74], among nurses was 13.47%, and among ancillary workers was 12.52% [69]. Several other studies [70, 75, 76] worldwide confirm the fact that the influenza vaccination coverage among physicians is the highest. In general, physicians are more prone to accept vaccination than other HCWs, e.g. the full hepatitis B vaccination coverage among physicians is 2.6 times higher than nurses [18]. Several reasons could be behind this observation such as greater misconceptions about vaccines among nurses and other HCWs, less fear and care about infectious diseases, less knowledge and more doubt about vaccine efficacy. This finding is a major concern in health care settings especially during the COVID-19 pandemic since nurses and assistant nurses have more and longer direct contact with patients than other HCWs [77]. Also, the seroprevalence of SARS-CoV-2 antibodies is higher among frontline health care workers and health care assistants [78] indicating that nurses and assistant nurses represent a high-risk group for SARS-CoV-2 infection.

We found that older age was related with an increase in willingness to get vaccinated against COVID-19. This finding is unsurprising since HCWs are quite familiar with the fact that older age is one of the strongest risk factors for COVID-19 mortality [79–81]. Therefore, it is more probable for older HCWs to take the COVID-19 vaccine due to their own self-interest. In a similar way, we found that HCWs with chronic conditions were more prone to get vaccinated against COVID-19. This finding makes sense since HCWs with comorbidity is a high-risk group for complications and death from COVID-19 as this is the case for the general population also according to several meta-analyses [80–84]. Older HCWs with comorbidity confront COVID-19 with fear and anxiety affecting critically their decision to accept a COVID-19 vaccine. An interesting result in our review is that male gender was associated with greater likelihood of taking COVID-19 vaccine. Two reviews regarding influenza vaccination [19] and hepatitis B vaccination [18] did not find any relation between gender and vaccination coverage. A possible explanation for our observation could be that the individual perceived risk about COVID-19 is higher among male HCWs.

According to our study, being vaccinated against flu during previous season was associated with COVID-19 vaccine acceptance. Similarly, HCWs with vaccine confidence and positive attitude towards a COVID-19 vaccine were more likely to be vaccinated against COVID-19. These findings are of utmost importance since the WHO named vaccine hesitancy as one of the top ten threats to global health in 2019 [85]. Health care workers especially at primary care should communicate in a clear way the message that vaccines are safe and effective to improve vaccination coverage in communities [86]. Since a safe and effective COVID-19 vaccine seems to be the only solution for this pandemic, the positive attitude of HCWs towards vaccination is imperative. Vaccine hesitancy among HCWs with regard to other vaccines, such as seasonal influenza vaccine already exists.[87–89] In case of the COVID-19 vaccine the situation can be worse since vaccine hesitancy is fuelled by fake news and conspiracy theories [90]. The reluctance or refusal of HCWs to vaccinate against COVID-19 could diminish the trust of individuals and trigger a ripple effect in the general public [91, 92]. There is a need to build confidence and trust in communities to rollout successfully a COVID-19 vaccine.

Additionally, we found that individual perceived risk about COVID-19 was related with increased COVID-19 vaccination acceptance among HCWs. HCWs may be reluctant to receive a novel COVID-19 vaccine when they believe that it is not protect against a significant personal threat. On the other hand, the self-perceived susceptibility to and seriousness of a vaccine infectious disease such as COVID-19 may increase vaccine acceptance [93]. This association has already observed in case of COVID-19 not only in the general public [94] but also in HCWs[48]. A warning sign to public health safety is that vaccine hesitancy is greater among nurses than among physicians [95–97].

Our study is subject to several limitations. In particular, more than the half of studies was of moderate quality, while four out of 24 studies were published in pre-print services which do not apply peer-review process. We performed subgroup analysis according to studies quality and publication type to overcome this limitation. The statistical heterogeneity in results was very high due probably to variability in study designs and populations. In that case, we applied a random effects model and we performed subgroup and meta-regression analysis. We included all studies conducted till to July 14, 2021 but vaccines are now available and HCWs attitudes towards COVID-19 vaccination could be changed for this reason. Our meta-regression analysis showed that the proportion of HCWs that intend to accept COVID-19 vaccination was independent of the data collection time but studies of current attitudes should be performed. Data with regards to the factors related with intention of HCWs to accept COVID-19 vaccination were limited, while five studies used multivariable models to eliminate confounding. We consider this as a potential area for future study. Moreover, all the studies included in this review were cross-sectional studies making causal inferences impossible. Finally, the proportion of HCWs that intend to accept COVID-19 vaccination may be an overestimation since studies evaluated self-reported answers that could be subject to social desirability bias, with HCWs knowing that the general public expects a high COVID-19 vaccination coverage among them.

## Conclusion

HCWs are identified worldwide as priority recipients of the novel COVID-19 vaccine since they represent a high-risk group for SARS-CoV-2 infection and transmission risk of SARS-CoV-2 in clinical settings between patients and HCWs is high. Also, HCWs serve as trusted community workers on public health topics and their role in promoting COVID-19 vaccine acceptance is critical. Thus, COVID-19 vaccine hesitancy among HCWs should be eliminated to inspire the general public towards a positive attitude regarding a novel COVID-19 vaccine. We found a great variability of COVID-19 vaccination acceptance among HCWs worldwide and knowledge of the factors that influence this acceptance would be essential to provide information about vaccination programs, determine priority groups for vaccination, take extra protective measures, etc. Knowledge of the factors that affect intention of HWCs to accept COVID-19 vaccination is limited and there is an urgent need for further studies to make more valid inferences. Since vaccination is a complex behavior, understanding the way that HCWs take the decision to accept or not COVID-19 vaccination will give us the opportunity to develop the appropriate interventions to increase COVID-19 vaccination uptake and promote vaccination programs worldwide.

## Data Availability

Data will be available after request

## Acknowledgments

none

## Funding

none

## Author contributions

P.G, D.F. and D.K. were responsible for the conception and design of the study. P.G, I.V., D.F., A.B., and D.K. were responsible for the acquisition, analysis and interpretation of data. All the authors drafted the article or revised it critically for important intellectual content.

---

**Supplementary Figure S1.**
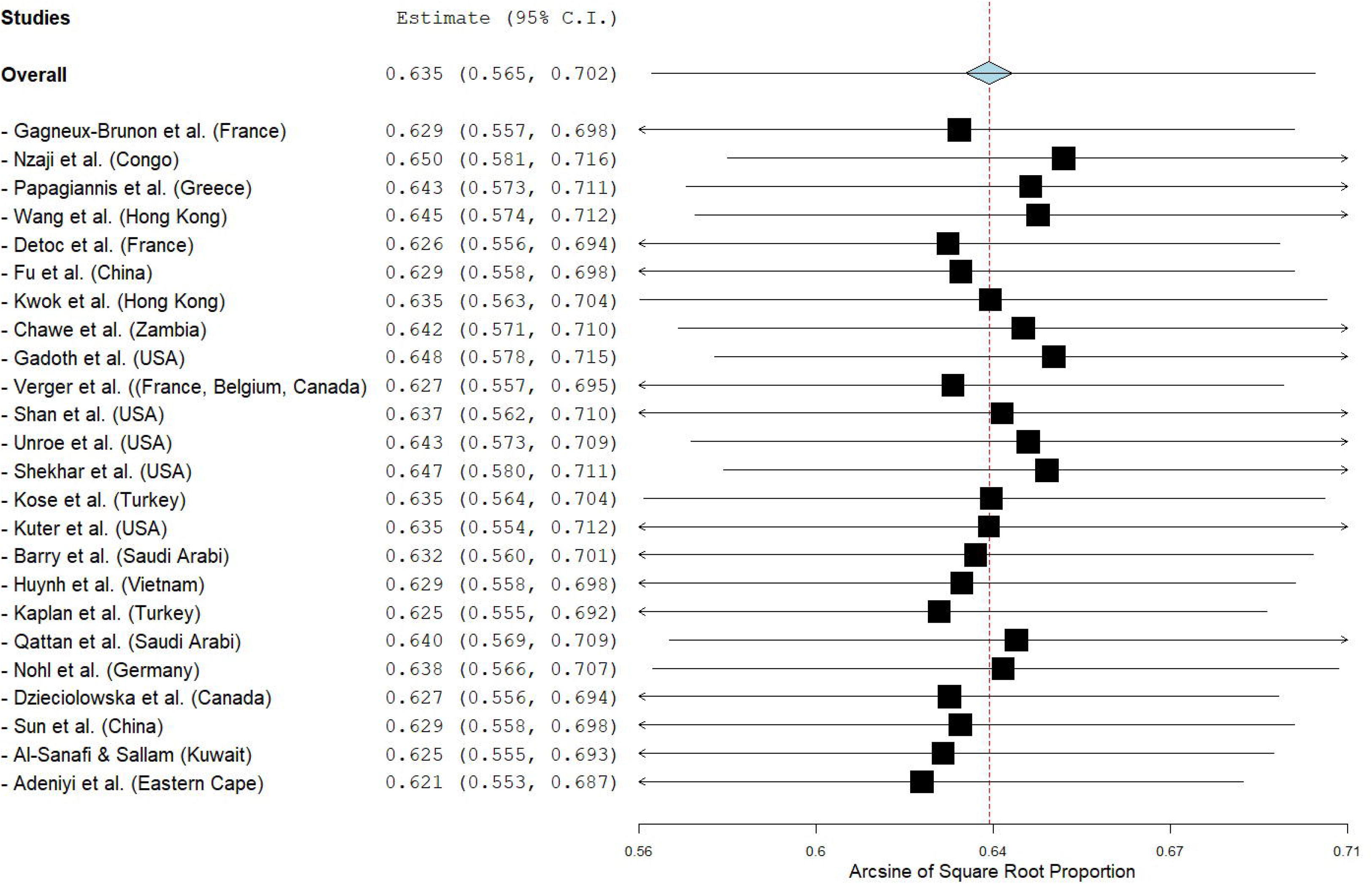
A leave-one-out sensitivity analysis of the proportion of HCWs that intent to accept COVID-19 vaccination.

## Notes

### Competing Interest Statement

The authors have declared no competing interest.

### Funding Statement

None to declare

### Author Declarations

No IRB/oversight body has to approve the study since it is a review

